# FEAT: A Flexible, Efficient and Accurate Test for COVID-19

**DOI:** 10.1101/2020.06.04.20122473

**Authors:** Linjiajie Fang, Bing-Yi Jing, Shen Ling, Qing Yang

**Affiliations:** Data Science and Analytics, Hong Kong University of Science and Technology; Mathematics, Hong Kong University of Science and Technology; Mathematics and Statistics, Dalhousie University, NS, Canada

**Keywords:** Epidemiology Control, Blood Testing, COVID-19, Testing Efficiency

## Abstract

Early detection of COVID-19 is critical in mitigating the spread of the virus. Commonly used tests include nucleic acid detection, antibodies detection via blood testing and CT imaging. Some tests are accurate but time-consuming, while others are cheaper but less accurate. Exactly which test to use is constrained by various considerations, such as availability, cost, accuracy and efficiency. In this paper, we propose a Flexible, Efficient and Accurate Test (FEAT). FEAT is based on group testing with simple but careful design by incorporating ideas such as close contact cliques and repeated tests. FEAT could dramatically improve the efficiency and/or accuracy for any existing test. For example, for accurate but slow test such as RT-PCR, FEAT can improve efficiency by multiple times without compromising accuracy. On the other hand, for fast but inaccurate tests, FEAT can sharply lower the false negative rates (FNR) and greatly increase efficiency. Theoretical justifications are provided. We point out some scenarios where the FEAT can be effectively employed.

## 1 Introduction

Population screening is essential for COVID-19 control. Commonly used tests for COVID-19 fall into three main categories: virus detection, antibodies detection via blood testing and CT image recognition. Reverse Transcription Polymerase Chain Reaction (RT-PCR) is one of the main testing methods for detecting coronavirus, due to its high accuracy and convenience. However, RT-PCR is a technical intensive testing method, which requires professional technicians to perform in some specialized laboratories [1]. As a result, RT-PCR is relatively expensive and time-consuming [2]. Its high cost and long testing time prevent it from widespread testing, particularly in less developed regions.

Efforts have been made around the world to have alternative diagnostic test kits in order to make the COVID-19 testing faster and more affordable. For example, in USA, Abbott developed a portable device “ID NOW”, which can detect positive results within 5 minutes, and negative results within 13 minutes [3]; in Bangladesh, a research group developed another fast and cheap test, known as “Dot blot test”, which reportedly costs only USD 3 and delivers results under 15 minutes, and the testing can be readily conducted in most labs in the country [4]. However, faster tests may not necessarily guarantee accuracy. In fact, there remains much concern about the high false negative rate (FNR) of these rapid tests. “*In Procop*’*s analysis, Abbott*’*s test had a false-negative rate of 14*.*8 percent*”, according to *The Scientists* [5]. The nearly 15% FNR would result in a large number of individuals released by mistake, which would have serious adverse effects.

Clearly, each diagnostic test has its own limitations, see Table 1 for a brief summary. Given these limitations, the natural question is whether it is possible to design some viable procedures to overcome the shortcomings of each diagnostic test. In our view, an ideal testing procedure should possess some, if not all, of the following metrics: accuracy, efficiency, flexibility and feasibility. The accuracy is measured by FNR and false positive rate (FPR). The efficiency is assessed by the expected number of tests per person. The flexibility requires the testing to be easily adjusted for different scenarios. The feasibility concerns the practical implementation of the strategy, such as being affordable and timely. A good testing should be accurate enough for infection control, be efficient so that it is economical and time-saving, be flexible for different regions, and be feasible to be implemented in reality.

**Table 1:** Pros and cons of three main tests. RT PCR is accurate and expensive [6]. ID NOW and Bangladesh 3 dollar fast and cheap tests, but less accurate [3] [4].

|  | Accuracy | Speed | Cost |
| --- | --- | --- | --- |
| RT-PCR | high | 4 -6 hours | high(USD 125) |
| ID NOW | low | 15 mins | medium |
| Dot blot test | low | 15 mins | low(USD 3) |

Our answer to the above-mentioned question is affirmative. In this paper, we propose some *Flexible, Efficient and Accurate Tests* (FEAT), which can achieve all the good metrics a testing strategy should possess. For instance, FEAT1 is designed for those tests with high specificity and sensitivity, for the purpose of improving the efficiency and maintaining accuracy; FEAT2 is proposed to improve the accuracy as the first priority, and increase the efficiency.

The rest of this paper is organized as follows. Section 2 includes a literature review of group testing and strategies for COVID-19 specifically. Section 3 introduces the methodology and basic techniques of FEAT strategies. Section 4 provides both theoretical and empirical results of FEAT. We compare the performances of FEAT1, FEAT2 and Dorfman’s group testing as well as individual one-by-one testing in our empirical study. Finally, a conclusion is provided.

## 2 Related Work

The implementation of group testing strategy could be traced back to Dorfman [7], who introduced an approach to economically test syphilis using grouped blood samples. Wolf [8] gave an overview of the early history following Dorfman’s original work. In formal terms of group testing problem, the items that cause a group to test positive are called *defective* items. And in our paper, we treat people who are infected with COVID-19 as *defective*.

In general, group testing could be classified by their testing design, the assumption of the diagnostic tests and the goal of detection.

- **Adaptive vs. Non-adaptive:** In the adaptive setting, the tests are processed sequentially, where the latter tests depend on the results from previous results. As for non-adaptive group testing, test pools are designed in advance (*design*) and the determination of defective items (*decoding* or *detection*) is given after all testing results are known. We consider adaptive group testing problem in our case, since the necessity of further tests is determined by the previous results.
- **Noiseless vs. Noisy testing:** In noiseless setting, diagnostic tests are assumed to work perfectly. However, we aim to tackle more realistic situations. And one of our contribution is exactly concerning the strategy of using unreliable diagnostic tests to give reliable results. Thus we only consider noisy testings.
- **Zero-error vs. Small-error:** Generally, the potential goal of group testing problems is either to recover all defective items (*zero-error*) or to recover the defective set with “high probability” (*small-error*). In reality, it is impossible to find out all infected people without any error by the unreliable tests. On the other hand, it is also hard to specify which “high probability” of recovering the set of infected people is acceptable. Thus we follow realistic demands: we use false negative rate and false positive rate as criterion. The goal of our group testing problem is to control the level of both false negative rates and false positive rates (such as being comparable to a single RT-PCR test), and at the same time to reduce the total number of tests.

Many algorithms have been developed to solve noiseless non-adaptive group testing problems, such as Combinatorial optimal matching pursuit (COMP), Definite defective algorithm (DD), Sequential COMP (SCOMP) and Smallest satisfying set (SSS) [9] [10]. And the aforementioned methods have their corresponding noisy versions, such as: noisy DD and noisy COMP [10]. Sterrett [11], Li [12] and Finucan [13] also proposed adaptive variants of the original Dorfman’s two-stage group testing. Binary-splitting algorithms [14] [15] is very efficient, yet it required too many testing stages even for small population, which is troublesome in process. Finuan [13] provided the theoretical proof of the optimal group size to achieve the best efficiency. De Bonis [16] proposed the estimation of minimum number of tests required for different number of stages with group testing. Patsula [17] compared the multi-stage grouping with some other adaptive testing approaches. Mcmahan [18] and Aprahamian [19] considered the heterogeneity of infections, which proposed a group testing strategy to test individuals based on the risk level.

By the high efficiency of group testing, it is considered an essential procedure to speed up COVID-19 detection. The implementation of group testing to detect COVID-19 was first suggested by researchers at Technion and Rambam Health Care Campus [20] [21] using RT-PCR. Nguyen [22] proposed the optimal pooling size based on the prevalence, with the consideration of disease progression and dilution of samples. Eberhardt [23] introduced a multi-stage grouping testing method under noiseless assumption without a discussion of the selection of the optimal size. Pouwels [24] discussed the necessity and potential improvements for group testing, yet no actions suggested further.

In our work, we propose a flexible multi-stage noisy adaptive group testing strategy to handle the practical population screening for COVID-19 infections. FEAT provides a flexible bunch of testing strategies catering for different prevalence levels, available virus detection methods and the demands for screening accuracy from place to place. Repeated tests are applied for the purpose of sensitivity control. Taking FEAT1 and FEAT2 as example, we explain how FEAT can balance the trade-off between accuracy and efficiency under different scenarios. Considering the heterogeneity nature of infections, close contact cliques (CCCs) are implemented. Additionally, FEAT considers at most three stages to guarantee feasibility. And operational delay in sequential setting can be shrunk to acceptable level when CCCs applied and stage number controlled. The detail and examples are discussed in Section 3.

## 3 Methodology

### 3.1 Motivations and Key Components of FEAT

The successful implementation of FEAT hinges on the four key components: group testing structure, repeated tests, close contact cliques and flexibility of performance parameters. The usage of three-stage group testing structure guarantees high efficiency; repeated testing hedges the loss of sensitivity of group testing methods and can further enhance accuracy; applying close contact cliques improves the testing efficiency by considering the heterogeneity of infections; the flexibility of performance parameters makes it possible to easily apply FEAT in different situations.

#### Group Testing

The improvement in efficiency of Dorfman’s group testing procedure is substantial. For example, if Dorfman’s procedure was applied to RT-PCR tests in USA, the overall efficiency would be boosted seven more times than the current practice (Fig.8). Similar performance is also observed for other diagnostic tests.

However, one very undesirable drawback with naive use of Dorfman’s procedure is that the FNR would be doubled: when Dorfman’s two-stage group testing is adopted, the probability that a positive sample been falsely diagnosed as negative is

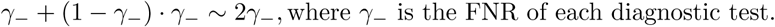

This problem is made even more pronounced when it is applied to tests with low sensitivity. For example, if the test kit has sensitivity level at 85%, or equivalently FNR at 15%, then Dorfman’s procedure would lead to overall 30% FNR. Intuitively, regarding to each single positive individual to be tested by Dorfman’s procedure, infections have twice chance to escape from being detected than individual tests. Similarly, despite the high efficiency, the direct use of N-stage group testing structure would amplify FNR to the order of *N* γ *γ*_−_, which should not be ignored under the noisy setting.

### Repeated Tests

To solve the increased FNR caused by group testing, we propose to use *repeated testing* procedure to either individuals or groups/subgroups level. When repeating the test once, we provide a positive result when there is either one test showing positive result. As a result, the false positives would be reduced significantly. For instance, if FNR = 15%, testing an individual twice would decrease FNR to 2.25%. A further theoretical discussion of FNR and FPR of FEAT is given in the *Appendix*.

Naturally, repeated tests would increase the total number of tests. The ensuing question is how to keep a balance between accuracy and efficiency, which is one of the focal points in this paper. We also provide flexible choices of FEAT1 and FEAT2 to encounter two situations where the accuracy of diagnostic tests are different.

### Close Contact Cliques (CCCs)

COVID-19 is a disease related to respiratory system. Contact with carriers is one of the necessary conditions for the spread of the virus [25]. It is observed that many infections occur within *close contact cliques* (CCCs), e.g., within family members, friends, colleagues, passengers from the same flights. Under these circumstances, when applying group testing, it would make sense to test CCCs together. Our results show the efficiency can be improved significantly. It is worth mentioning that FEAT performs very well even without using information of CCCs (see Fig.7).

### Feasibility and Flexibility

Feasibility and flexibility must be factored in any test strategy. In FEAT, we try to keep it as simple as possible so that it can be easily implemented in practice. Many existing designs are simply not feasible at all. For instance, the binary splitting method has been toyed around in the literature to minimize total number of tests [26], where individuals or groups, when tested positive, are further divided into equal halves, and so on. As a result, some individuals might need to undergo many rounds of tests, which we regard as unacceptable in practice. Along similar lines, when we employ repeated tests, we only need to require individuals to provide two independent samples at the same time. Such practice is very much standard in, say, providing urine samples for athletes in sporting competitions. Candidates are only required to gather A and B samples, then the repetition of gathering procedure is at most three times.

Another advantage of FEAT is its flexibility. Besides FEAT1 and FEAT2, users can choose any number of repetitions in each step by three parameters FEAT(a,b,c) with each number corresponding to the number of test repetition at each stage. In our case, FEAT1 and FEAT2 are the same as FEAT(1,2,2) and FEAT(2,2,2) respectively. The flexible choice of group size, i.e. the number of CCCs pooled in a group, can be adjusted according to the specific prevalence rate and the average size of such CCCs to achieve the best efficiency.

### 3.2 Methods

#### 3.2.1 FEAT1 for tests with high accuracy and low efficiency

For diagnostic tests with high accuracy but low efficiency, such as RT-PCR, we propose FEAT1 to largely improve the efficiency, see Fig.2 for illustration. We first form small subgroups, which are chosen as CCCs if available (see details in Section 3.2.3). Examples of CCCs include close family members, close neighbors, passengers from the same flight, etc. These subgroups are then pooled together to form larger groups. Tests will be performed for each group. Testing stops at this first stage only when the group is tested negative; otherwise it will move to the second stage, where each subgroup will be tested twice. Testing stops at the second stage only when the subgroup is tested negative in both occasions (- -); otherwise testing continues into the third stage where each individual will be tested twice. Again, testing stops only when the individual is tested negative in both tests (- -). For simplicity, we will refer to this test strategy as FEAT1.

**Figure 1:**
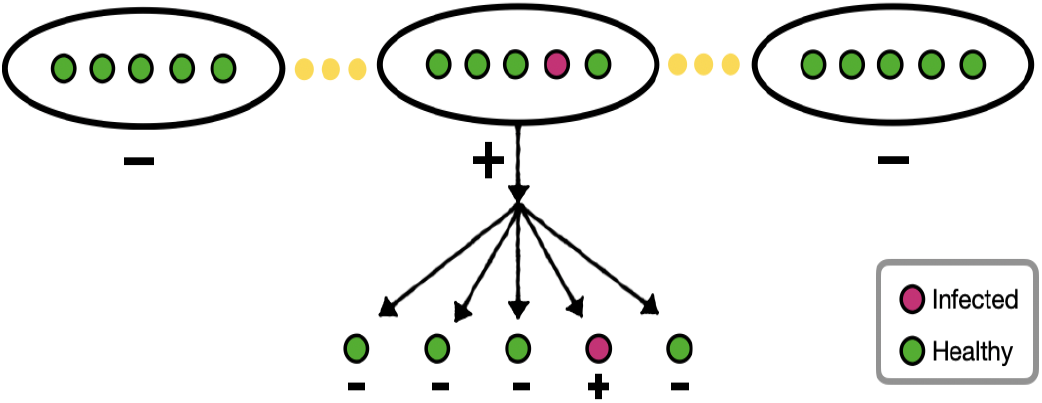
Dorfman’s two-stage group testing. In this graph, the pooling size is five (m =5). The groups of samples are tested and positive groups are later tested individually.

**Figure 2:**
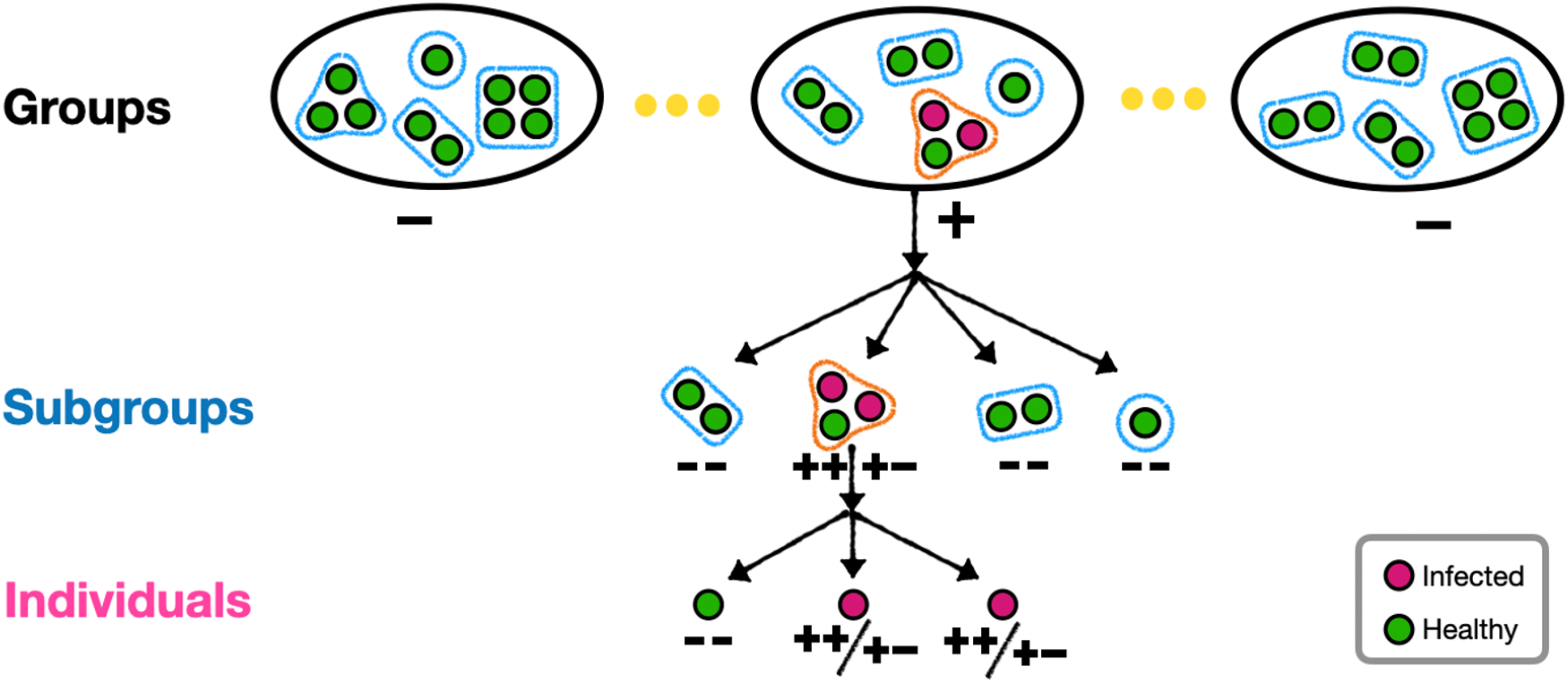
An illustration of FEAT1. Each group contains four CCCs (*k*^*\**^ = 4, where the subgroup sizes *m* ∈ {1, 2, 3, 4}). For group-level tests, FEAT1 performs one diagnostic test and stops the screening for negative groups. Subsequently, FEAT1 performs two diagnostic tests for each test simultaneously, and either one of the two diagnostic tests shows positive, such subgroup or individual would be labeled positive.

#### 3.2.2 FEAT2 for tests with high efficiency and low accuracy

When the sensitivity of available diagnostic tests is relatively low (e.g. 85%), one can consider some variants of FEAT1. For instance, we could test the groups twice in the first stage (Fig.3), and testing stops only when both testing results are negative; otherwise testing continues into the second stage, where the procedures are the same as in FEAT1. For simplicity, we will refer to this test strategy as FEAT2. As we will see later, FEAT2 can work extremely well for low sensitivity test kits.

**Figure 3:**
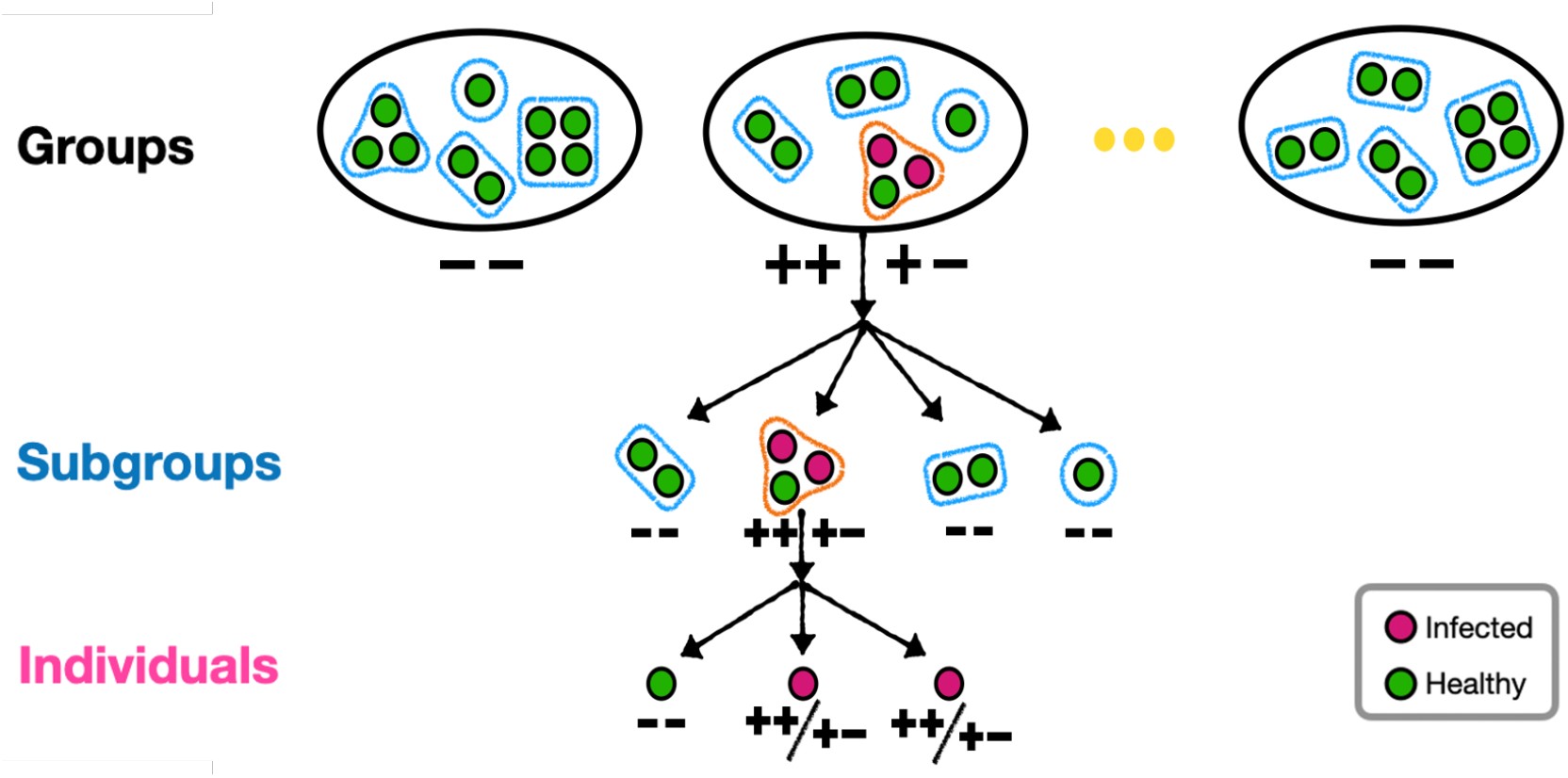
An illustration of FEAT2. Each group containing four CCCs (*k*^*\**^ = 4, where the subgroup sizes *m* ∈ {1, 2, 3, 4}). For group-level tests, FEAT2 performs two diagnostic tests and stops the screening for two negative results. Subsequently, FEAT2 performs two diagnostic tests for each subgroup/individual simultaneously, and either one of the two tests shows positive, such subgroup or individual would be labeled positive.

## 4 Theoretical Results

This section provides theoretical results of FEAT. Proofs of theorems are given in the *Appendix*.

### 4.1 Notations

The notations used in the following sections are summarized as below:

*ρ* : prevalence of the whole population.

*β* : probability that an individual is infected by the infected source in the same subgroup.

*k*^*\**^ : number of subgroups in a group in three-stage group testing. See examples in Fig.2, Fig.3. *m* : size of a subgroup.

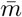 : average size of subgroups.

*k* : average size of groups. It satisfies *k* = *m* · *k*^*\**^.

*ρ*^*\**^ : fraction of infected subgroups. Also, 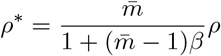, (see Appendix)

*γ*_−_ : FNR of each diagnostic test.

*γ*_+_ : FPR of each diagnostic test.

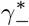 : FNR of the overall group testing.

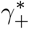: FPR of the overall group testing.

### 4.2 Optimal Pooling Sizes

The optimal number of subgroups in each group (see Fig.2, Fig.3) is closely related to the testing efficiency (see Fig.4, Fig.5). We extend the relationship between the group size and the prevalence level of two-stage methods in Finucan (1964) [13] to FEAT1 and FEAT2 respectively. The subgroups are naturally determined by clusters of people. For example, to screen a community, the subgroups are simply families; to screen travelers in the airports, the subgroups are people having close seat positions. We aim to find the number of subgroups per group to maximize the expected number of persons per test, i.e., to maximize the testing efficiency.

We refer the number of subgroups *k*^*\**^ that maximizes the efficiency as “optimal”, and denote that number as 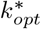.

#### Theorem 4.1

*For FEAT1, given the prevalence of subgroups ρ*^*\**^, *the optimal number of subgroups per group* 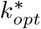 *is*

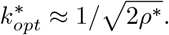

In our simulation (Fig. 4), we assume 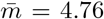 [27], *β* = 0.7, *ρ* = 0.005, thus *ρ*^*\**^ ≈ 0.006. We can quickly compute 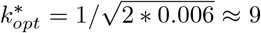, equivalently to pool 9 * 4.76 ≈ 43 people in a group, which is the same as our simulation result.

**Figure 4:**
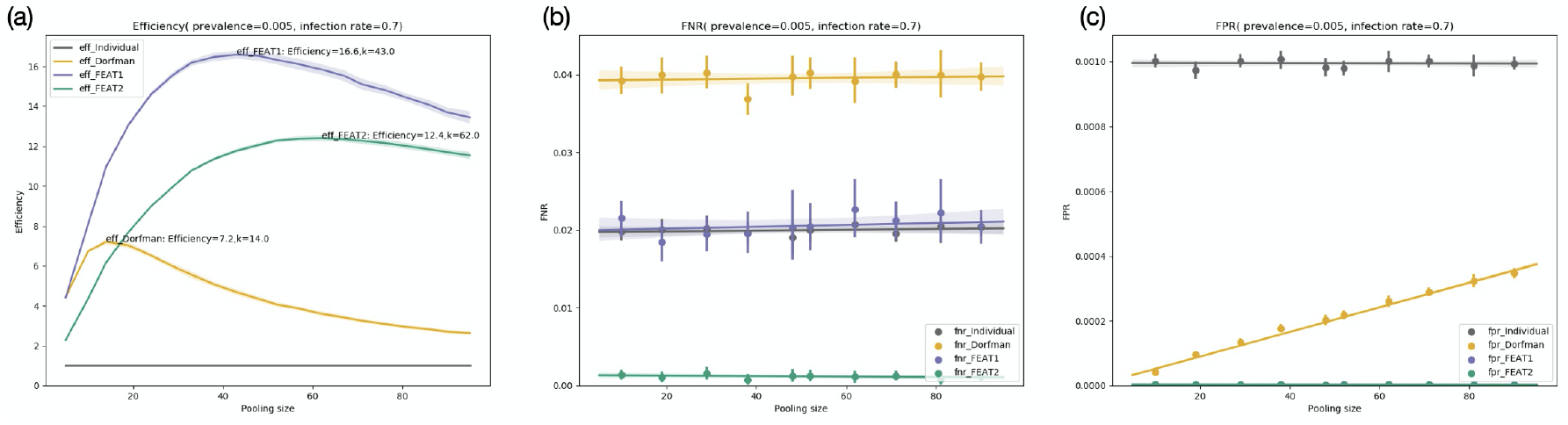
Comparisons of FEAT1, FEAT2, Dorfman’s method and the individual test on 50 simulation sets. Each simulation set contains 20,000 families with the distribution of household sizes following data in Bangladesh [29]. The FNR and the FPR of each single test are supposed to be 0.02 and 0.001 respectively. We assume the individual prevalence *ρ* = 0.005 and the infection rate within close contact clique *β* = 0.7. We evaluate the efficiency and accuracy along different pooling sizes. (a): The efficiencies versus pooling sizes. (b): The FNR versus pooling sizes. (c): The FPR versus pooling sizes. The FPR of both FEAT1 and FEAT2 overlaps each other.

#### Theorem 4.2

*For FEAT2, the optimal number of subgroups per group* 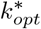 *is*

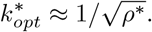

Under the same setting in FEAT1, 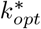 of FEAT2 is estimated by 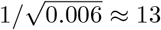, or equivalently to pool 63 people in a group, which is a little different from the simulation results in Fig.5. Fig.5 shows that it is robust for FEAT2 to apply group size that differs one or two subgroups from the optimal while maintain almost the best efficiency.

**Figure 5:**
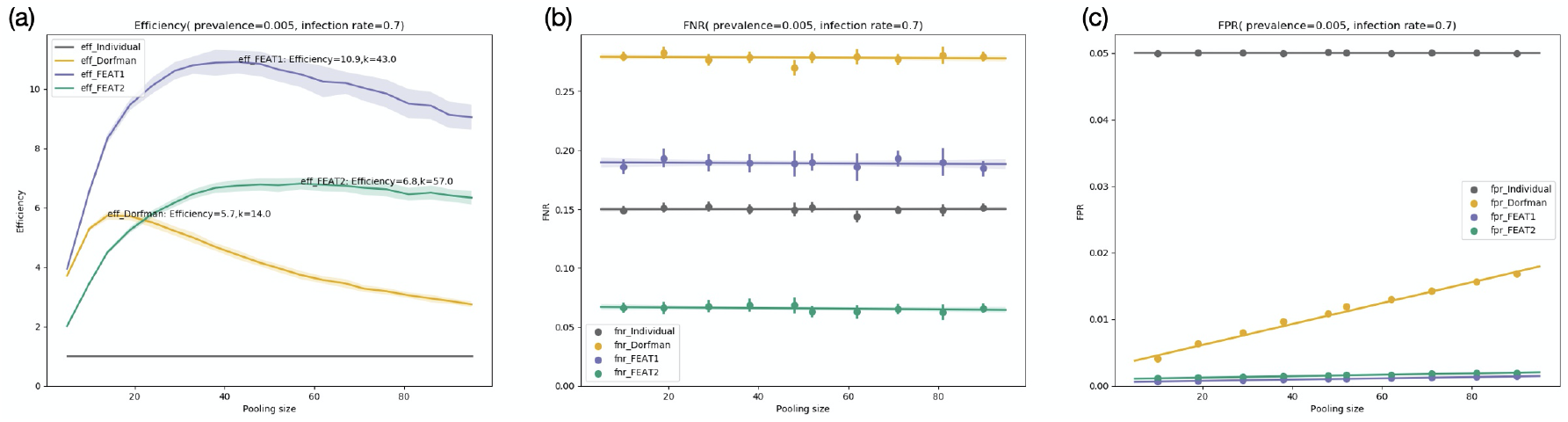
Comparisons of FEAT1, FEAT2, Dorfman’s method and the individual test on 50 simulation sets. The configuration is the same as Fig.4 except that the FPR and FNR are assumed to be 0.15 and 0.05 respectively. (a): The efficiency versus pool size. (b): The FNR versus pool size. (c): The FPR versus pool size.

### 4.3 Prevalence Condition for Efficiency

For Dorfman’s two-stage group testing, it has been shown that it is better than individual testing if and only if *ρ* < 1 — 1*/*3^1*/*3^ ≈ 0.306 [28]. It is natural to ask under which conditions the FEAT strategy is more efficient than testing individually. Here we give the results:

#### Theorem 4.3

*Suppose the diagnostic tests are perfect and the population is large enough. Then*,

- *FEAT1 is more efficient than individual testing if and only if:*

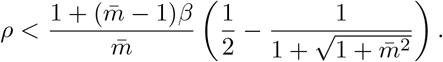
- *FEAT2 is more efficient than individual testing if and only if:*

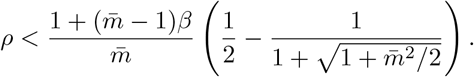

The upper bound for the prevalence is determined by the size of subgroups 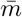 as well as the infection rate *β*. As illustration, if 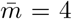 and *β* = 0.7, FEAT1 requires *ρ* < 0.24 and FEAT2 requires *ρ* < 0.19 to be efficient. If 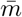 is changed to 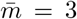, such requirements is changed to *ρ* < 0.21 and *ρ* < 0.16 respectively.

### 4.4 Accuracy of FEAT

We analyse the accuracy of FEAT by the overall FNR 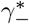 and FPR 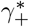

#### Theorem 4.4

*Suppose ρ is small, the order of the accuracy of FEAT is given by:*

- *For FEAT1:*

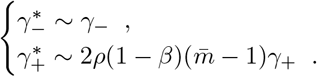
- *For FEAT2:*

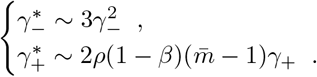

FEAT1 maintains the same order of FNR as individual testing while FEAT2 lowers the FNR to the second order. Although FEAT1 does not reduce FNR as FEAT2, FEAT1 is much more efficient without performing repeated testing in the first stage. The choice of FEAT1, FEAT2 or even other variants depends on the accuracy of diagnostic tests.

## 5 Empirical Results

To evaluate the performance of FEAT, we first perform empirical study of FEAT1 and FEAT2 for data of Bangladesh. In our study, we try to use real data as much as possible, such as: the prevalence, household distribution and accuracy of diagnostic tests. For other parameters. we make reasonable assumptions followed by sensitivity analysis. Finally, FEAT is implemented of FEAT for five countries, and compared with Dorfman’s group testing.

### Numerical Setup

In Fig.4 and Fig.5, we assume prevalence *ρ* = 0.005 and household distribution of Bangladesh is based on data [29]. That is, we choose the number of family members from {1, 3, 5, 7} with probabilities equal to {0.02, 0.30, 0.45, 0.23} respectively, and the average family size is 4.76. We also assume the infection rate within close contact cliques to be *β* = 0.7.

We first generate 20,000 families (as subgroups) with the fraction of infection equal to 0.006. Then we generate the number of members and infections of each family by the household distribution and the infection rate to make sure the overall individual prevalence is 0.005. We use pooling size, referring to the number of individuals in each group, as the changing performance parameter in comparing performances of different methods (Fig.4, Fig.5). For each pooling size, we simulate 50 replicates and evaluate the average performance.

In Fig.7, we follow the same configuration as in Fig.4 except for changing the infection rate *β* from 0 to 1 with a gap 0.1. For each *β*, we evaluate performances of each method by their optimal pooling sizes. Similarly, in Fig.6 we fix *β* = 0.7 and change prevalence from 10^−4^ to 10^−2^ with a gap of log-scale equal to 0.2 in order to evaluate the best performance on different prevalence levels. To avoid the variance when searching for the best efficiency, we simulate 30 replicates of simulation set containing 20,000 families and the average performance is compared.

**Figure 6:**
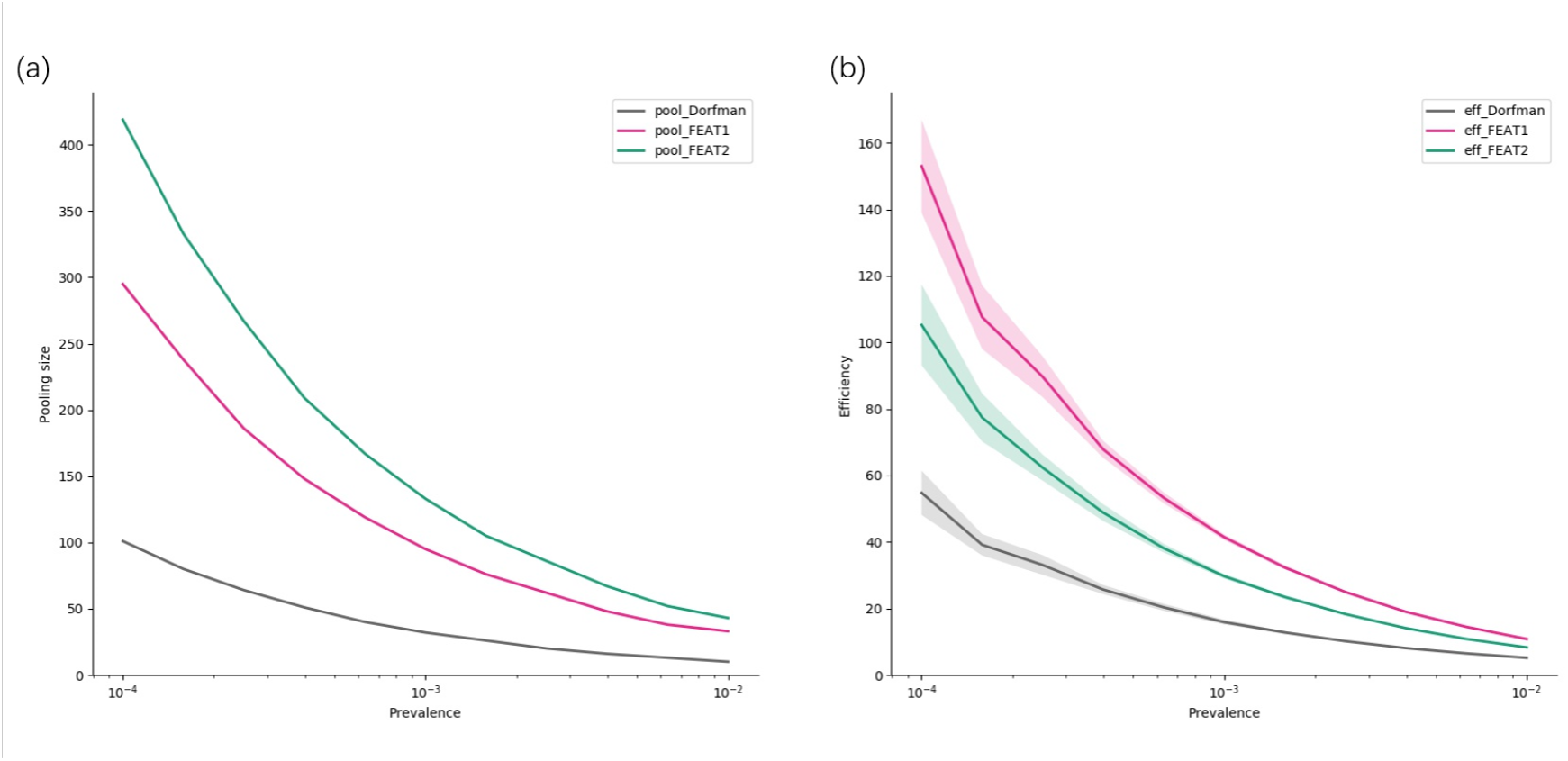
The best performance of FEAT1, FEAT2 and Dorfman’s method under different prevalence. The infection rate is fixed to be the same as Fig.4.(a): The optimal pooling sizes of three methods versus different prevalence. (b): The efficiencies of three methods versus different prevalence.

**Figure 7:**
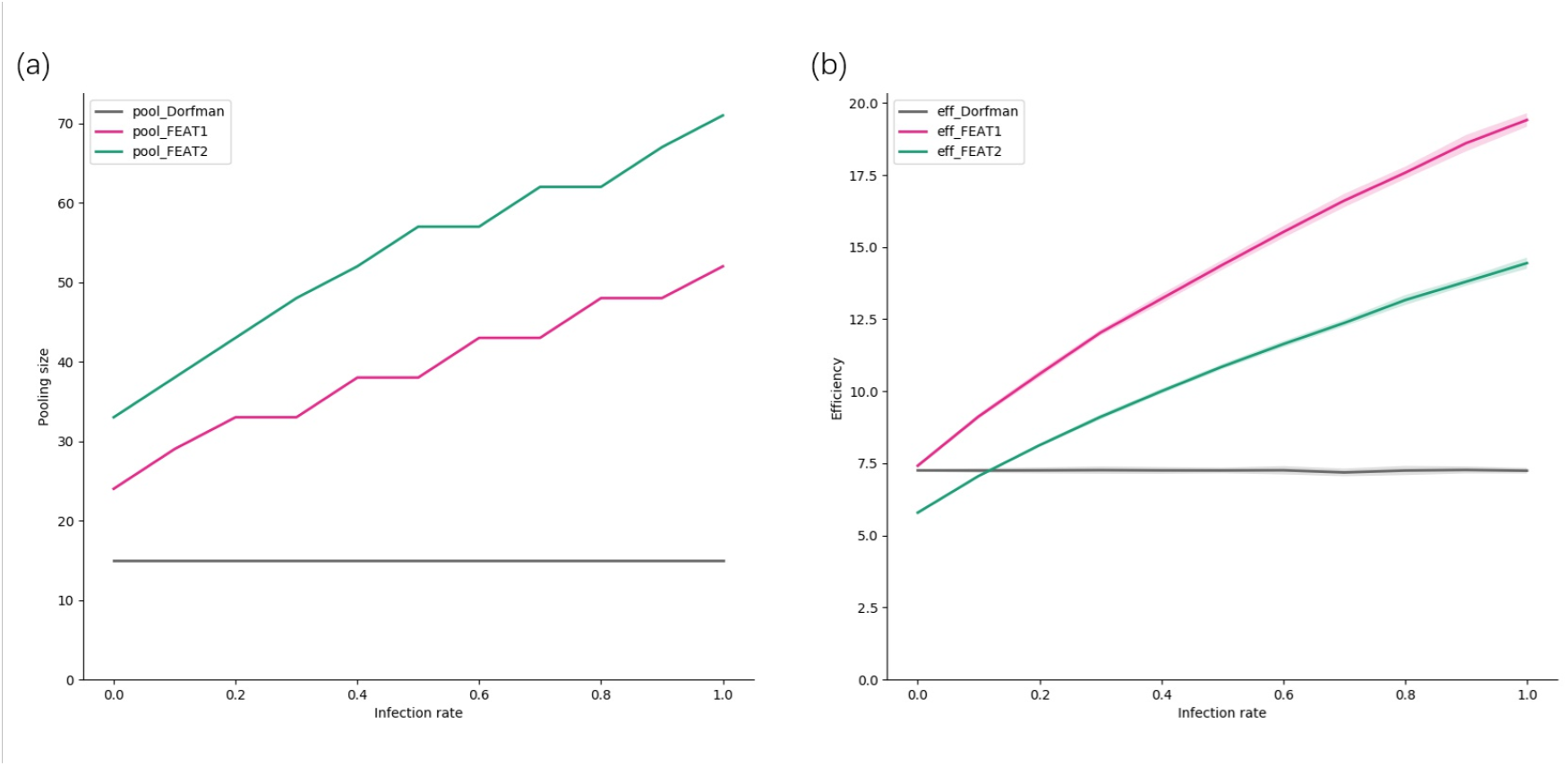
The best performance of FEAT1, FEAT2 and Dorfman’s strategy under different infection rate within close contact clique. The prevalence is fixed to be the same as Fig.4. (a): The optimal pooling sizes versus infection rate. (b): The best efficiencies versus infection rate.

### Results for Testing with High Specificity and Sensitivity

Fig.4 shows the results when accurate tests, such as RT-PCR, are used. We assume the FNR = 0.02 and FPR=0.001. We simulate our data following the household distribution of Bangladesh under prevalence rate 0.005, the results in other places are similar and the optimal performances are summarized in Fig.8, will be discussed later. Thus we make the following observations.

**Figure 8:**
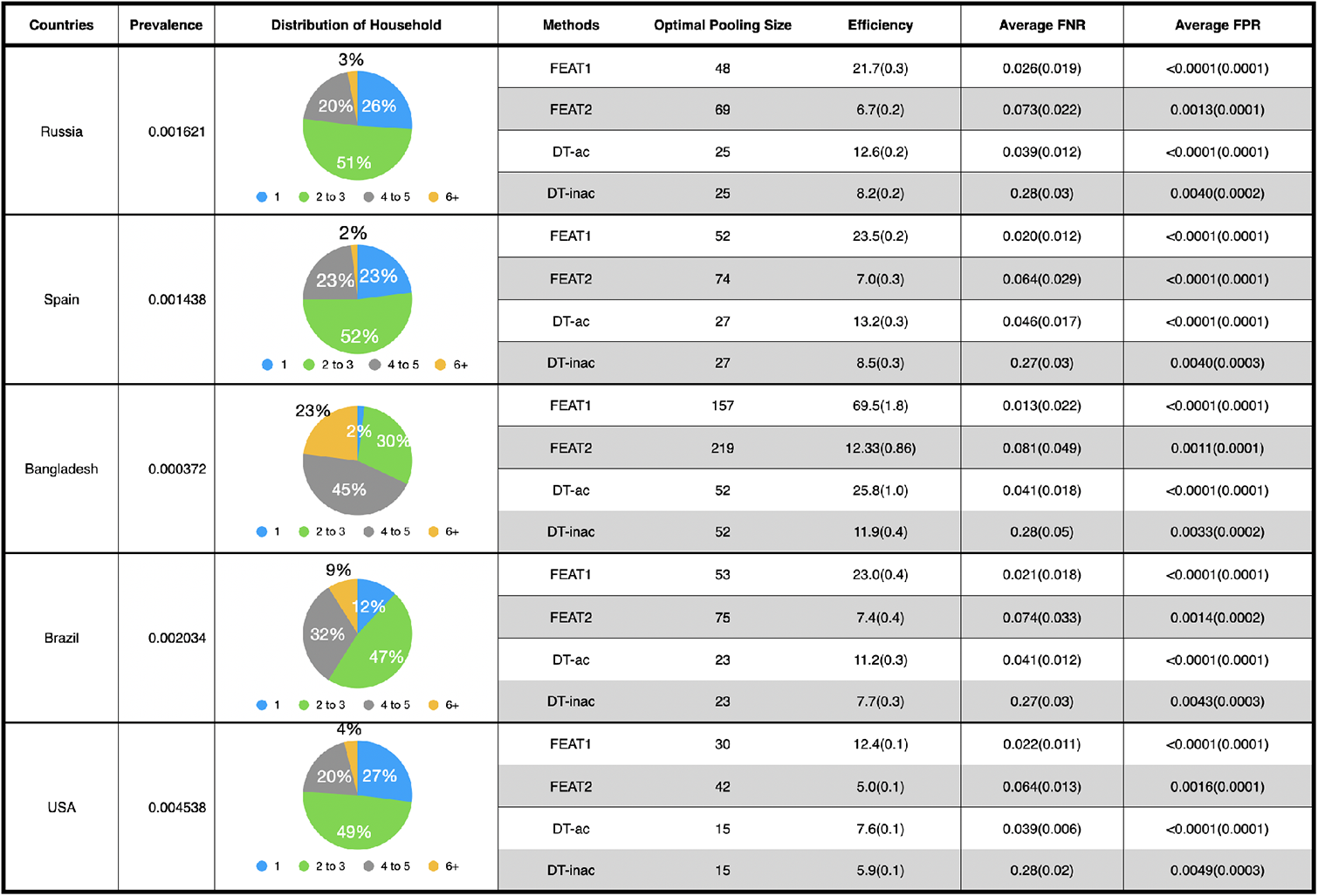
This table provides the FEAT and Dorfman’s two-stage group testing (DT-ac/DT-inac) results for five countries: Russia, Spain, Bangladesh, Brazil and United States of America (USA). The quantities are given by mean values with standard deviation in brackets. FEAT1 and DT-ac use accurate diagnostic tests(FNR =0.02, FPR=0.001). The FEAT2 and DT-inac use cheap but inaccurate diagnostic tests (FNR =0.15, FPR=0.05), whose rows are shadowed to differ from methods with accurate diagnostic tests. The infection rate within close contact cliques is supposed to be 0.7.

- FEAT1 and FEAT2 are over 16 and 12 times efficient as the individual test with optimal pooling sizes to be 43 and 62 respectively, as shown in Fig.4(a). The maximum efficiency of Dorfman’s method is 7.2, which is far less than FEAT. Here efficiency is measured by the average number of persons implemented for each single test.
- In terms of the accuracy, Dorfman’s method doubles the FNR from 0.02 to 0.04. FEAT1 nicely controls the FNR to be the same level of the individual test. Although FEAT2 is less effective than FEAT1, the FNR of FEAT2 is extremely small, see Fig.4(b).
- The FPRs of FEAT1 and FEAT2 are nearly zero and overlap each other, and the FPR of Dorfman’s method is almost linear in pooling size. Fig.4(c) shows that the FPR of all the group testing methods are far less than the individual test.
- Overall, FEAT1 could improve efficiency remarkably and at the mean time maintain the high accuracy.

### Results for Testing with Lower Specificity and Sensitivity

If RT-PCR is not available or not affordable in some nations or regions, we would consider testing strategies applying only cheap but inaccurate tests. Fig.5 shows the performances of group testing strategies applying inaccurate tests. The configuration of the simulation is the same as above except that we assume FNR=0.15 and FPR=0.05. We make following remarks.

- FEAT1, FEAT2 and Dorfman’s method reach maximum efficiency 10.9, 6.8 and 5.7 respectively, refering to Fig.5(a). It is noticed that efficiencies under inaccurate tests are less than the scenario when accurate tests are applied, as higher FPR (0.05 v.s. 0.001) could lead to redundant tests on groups with no infection.
- From Fig.5(b). Dorfman’s method even doubles the FNR from 0.15 to 0.30, which is unacceptable. FEAT1 could no longer control the FNR to be the same as the individual test in such scenario, which is increased to be around 0.2.
- FEAT2 reduces the FNR substantially from 0.15 to 0.0675. In terms of the FPR, Fig.5(c) shows that FEAT have consistently better sensitivity than Dorfman’s method.
- FEAT2 can leverage cheap inaccurate tests and largely reduces the overall FNR. For example, if FNR=0.10 for the diagnostic test, then FEAT2 can reduce the FNR to 0.03; and if FNR=0.05, FEAT2 can reduce it to 0.0075. Details of the calculation are given in *Appendix*.

### Effects of the Prevalence

Fig.6 compares the performance of FEAT1, FEAT2 and Dorfman’s method under different prevalence levels. To control the number of parameters, we fix the infection rate within close contact clique *β* = 0.7 and change the prevalence from 0.01% to 1%. Fig.6(a) shows that optimal pooling sizes of all group testing methods decreases when the prevalence increases. Fig.6(b) shows that the efficiencies of three methods also decrease with prevalence. The smaller the prevalence, the more efficient the group testings should be.

### Effects of the Infection Rate within Subgroups

Fig.7 compares the performance of FEAT1, FEAT2 and Dorfman’s method with optimal pooling sizes under different infection rates *β* ∈ [0, 1]. Here we fix the prevalence *ρ* = 0.005. Fig.7(a) shows that FEAT methods always pool more samples than Dorfman’s method and the pooling size is monotonously increasing with respect to *β*. Fig.7(b) shows that when *β* ≥ 0.15, FEAT2 would be more efficient than Dorfman’s method, and FEAT1 always performs better than Dorfman’s regardless of *β*. Since SARS-CoV-2 spreads very easily and sustainable between people [30], it is reasonable to speculate that FEAT would be more efficient in reality.

### Implementation of FEAT

In Fig.8, we evaluate the optimal performances of FEAT in different countries. The number of confirmed cases, death and recovered (data collected from [31] before June, 2020) and the population [32] are used to calculate prevalence in each country. The distribution of households is gathered from United Nations reports in 2017 [29]. For simplicity, here we compare FEAT1 equipped with accurate diagnostic tests (FNR =0.02, FPR=0.001), FEAT2 equipped with inaccurate diagnostic tests (FNR=0.15, FPR=0.05), Dorfman’s with accurate tests (FNR =0.02, FPR=0.001, denoted by DT-ac) and Dorfman’s with inaccurate tests (FNR =0.15, FPR=0.05, denoted by DT-inac). Group testing methods with cheap but inaccurate tests are shadowed.

Fig.8 provides FEAT solutions for five countries whose number of confirmed cases is large. Comparing FEAT1 with DT-ac, The accuracy of all five countries are well controlled, where FNR is around 0.02 and FPR is less than 0.0001. In terms of efficiency, FEAT1 surpasses DT-ac, and averagely doubles the efficiency. Additionally, in some countries with larger average family size such as Bangladesh, the efficiency of FEAT2 is as good as DT-inac. However, its accuracy can be largely improved.

## 6 DISCUSSION

### How Accurate Can Group Testing Achieve?

If repetition is not adopted, three-stage group testing, which applies to the groups of subgroups, is much more efficient than two-stage grouping. However, compared with individual tests, two-stage grouping (like Dorfman’s method) doubles FNR, and its FPR is nearly linear to the group size. Three-stage grouping triples FNR, and their FPR is nearly linear to the subgroup size. These facts direct us to avoid overmuch number of stages and prevent pooling too many individual samples in the last stage of group testing strategy.

To alleviate the problem of multiplying FNR without loss of much efficiency, FEAT adopts a three-stage grouping structure and performs repeated tests at proposed levels, whose FNR is controlled to be at the level of second order and FPR is stable along different pooling sizes. Although repetition intuitively increases the total number of tests, by virtue of three-stage structure and infection information, FEAT maintains comparable efficiency as Dorfman’s method.

### Necessity of Group Testing

When the prevalence becomes very high, both efficiency and accuracy of group testing will be greatly reduced. Thus we have to consider if it is necessary to implement group testing when the level of prevalence is severe. Fig.9 provides theoretical curves of naive two-stage grouping testing with repeated tests, which suggests that when prevalence is over 10%, the number of tests of grouping patients would outnumber the individual tests. It could be a loss of the original intention for designing group testing. For areas with very severe epidemics, small pooling size will be appreciated or just remain as simple as individual testing. However, to the best of our knowledge, prevalence in most areas for COVID-19 is about 1% [33] or even lower, so the necessity of group testing is still evident for COVID-19 pandemic.

**Figure 9:**
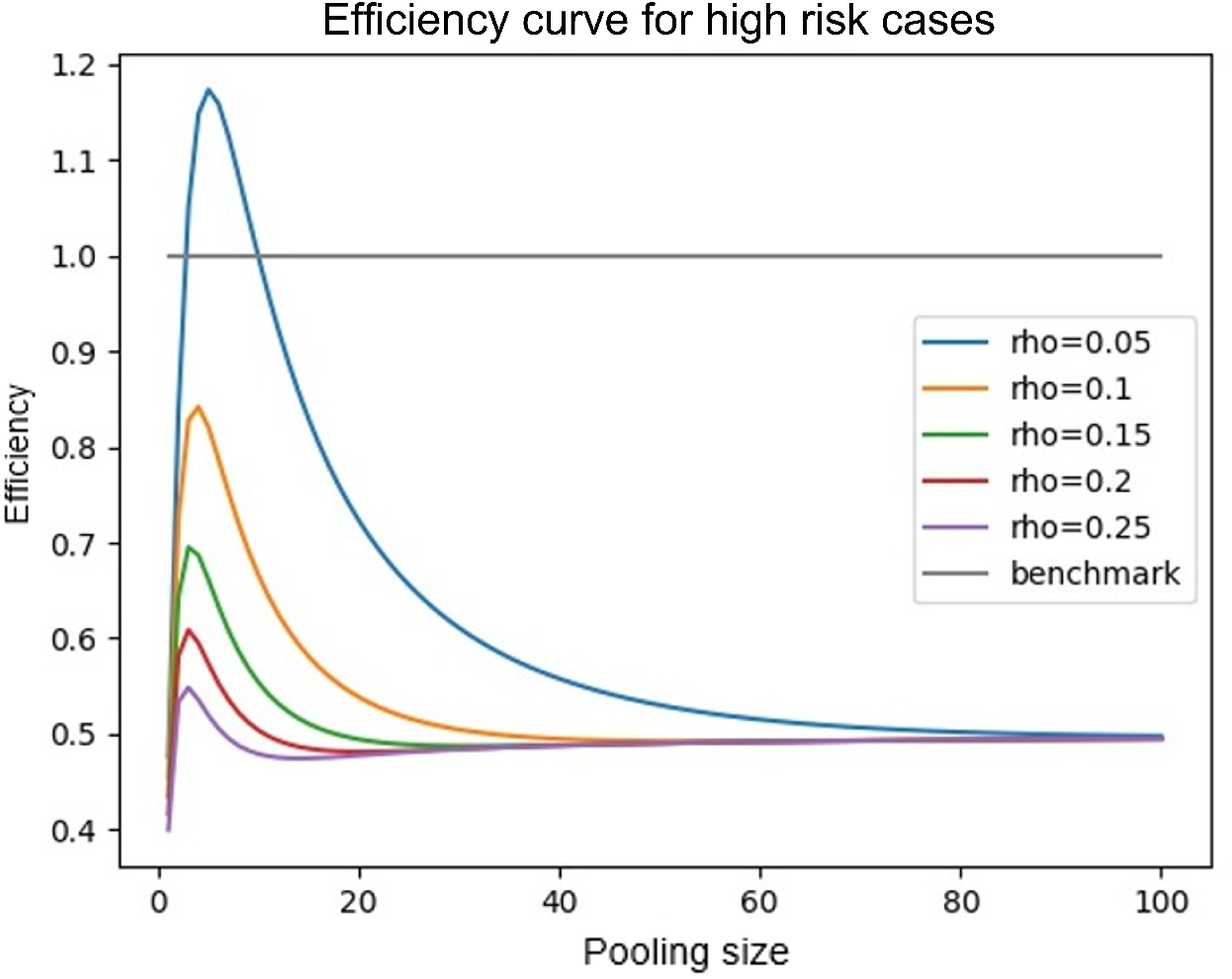
The efficiency of two-stage group testing method with repeated tests when the prevalence level is high.

### Determination of Close Contact Cliques

The concept of close contact cliques can be freely generalized to any clique of individuals with close contacts, not only families. For example, the residential relations, roommates staying in the same dormitory unit. Furthermore, when applying group testing strategies in airport to diagnose the virus status of commuters, passengers sitting in front and back rows on the same plane can be pooled in the same close contact clique.

Unlike Dorfman’s method, which pools a fixed number of individuals, FEAT pools samples by a fixed number of close contact cliques. Since the size of close contact cliques is flexible, FEAT pools different number of samples from group to group, which concerns more about the contact information than restrictions on specific pooling size. In this paper, the contact between groups or subgroups is ignored. However, in reality, the contact between people is much more complicated. The contact information of patients plays an important role to distinguish high risk susceptible patients from low risk ones. A flexible grouping strategy based on contact network might help cluster the infected patients to certain pools and largely reduce the number of tests.

### Potential Operational Delay

Group testing is a sequential testing strategy, where operational delay could be a natural concern. In this part, we would explain there should be few individuals delayed by three-stage structure. And under current quarantine, delay will not lead to any inconvenience in some situations.

First, compared to some multi-stage group testing methods, FEAT allows at most two times delay from individual tests. Fig.10 shows the operational delay of FEAT when *ρ* = 0.005 and *k* = 32. Only less than 1 percentage of the whole population would be required to take the third-stage test. However, binary splitting for group tests delays four times when the population size is only 30. On the other hand, FEAT is a set of methods introduced for testing strategies, where users also can implement two-stage structure to reduce the total operational delay.

**Figure 10:**
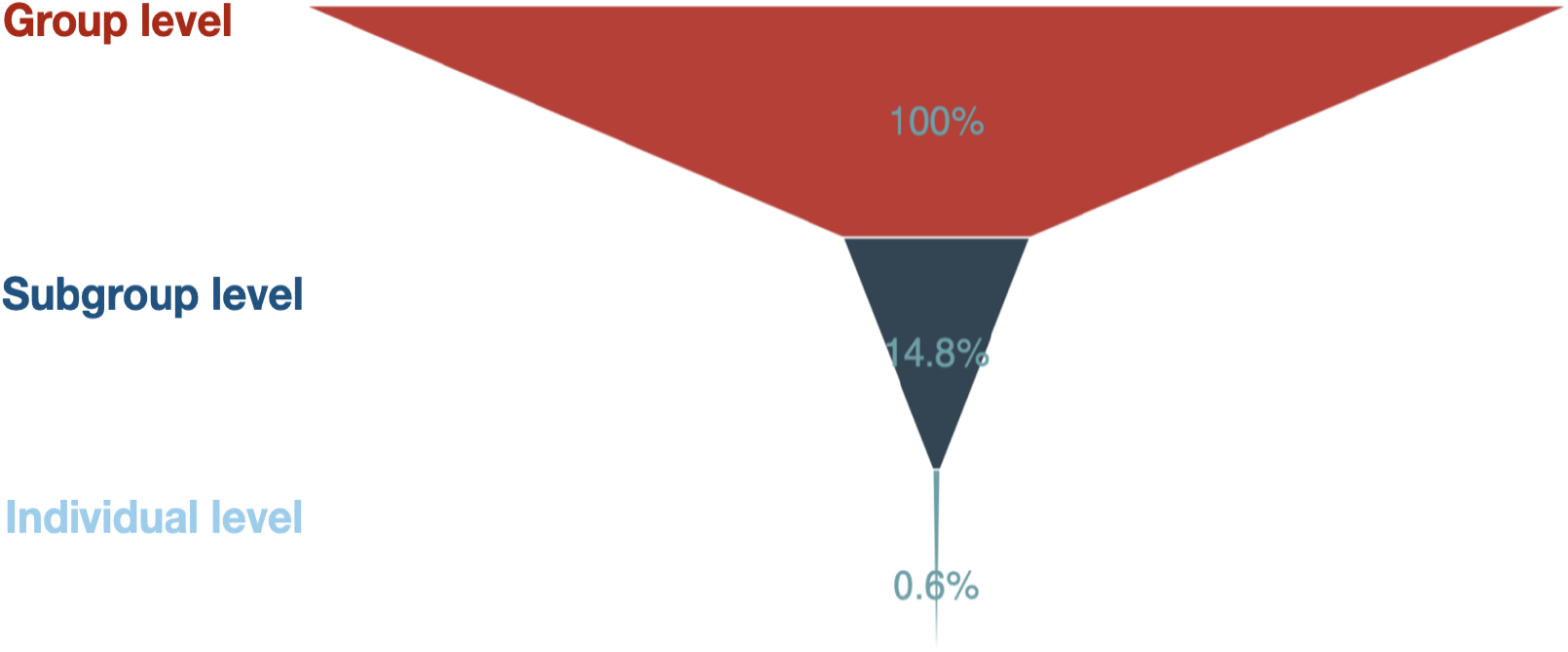
The operational delay of FEAT. We assume prevalence *ρ* = 0.005, group size *k* = 32 and the diagnostic tests are perfect. Only a very small fraction of people (< 1%) will take all three stage tests.

Second, under current quarantine policy, international travellers are mandatory to take at least 2-week quarantine in the hotel when access to China. In this situation, three-stage group testing does no harm, as infected cases have no chance to contact with others before confirmed.

Next, FEAT as well as other group testing based strategies is design for population screening, instead of using with high-risk cohorts or in clinical settings to conclude the viral status of potential patients who already have symptoms. For that reason, the overall prevalence of population should not be high. With the low prevalence, the majority of population should be screened out as negative at the first stage of group testing, where there would be no operational delay.

### “Predictive” Group Testing

In some areas, government would propose universal testing among all the population, where millions of individuals could be involved. In order to perform a more efficient testing, it is prioritized to test people who have high probability of getting infected. Before testing, we can make prediction of the risk level for someone based on their contact information, symptoms and residential areas. Such risk prediction is usually non-uniform among different groups of people. In the future, we hope to develop methods to predict the risk of people and build testing strategies based on different predictions.

## 7 Conclusion

We proposed the FEAT procedures by incorporating several ideas such as group testing, close contact cliques (CCCs) and repeated tests. Different versions of FEAT can be designed to cater for different scenarios. We designed two such versions, FEAT1 and FEAT2, corresponding to several different diagnostic testing kits. FEAT can increase the overall testing efficiency and accuracy. The improvement in efficiency can reduce the number of kits required for a specific size of population. In other words, the financial burden for testing will be reduced significantly. Additionally, the contribution of increasing accuracy is also meaningful. Highly accurate test methods can be applied in some areas when those accurate tests are not available. In terms of testing flexibility, different versions of FEAT (e.g. FEAT1 and FEAT2) can cater for different situations to balance both accuracy and efficiency. Finally, the mechanisms and application in countries are described in detail in this paper.

## Data Availability

No real data applied, simulation is included in the link.

## 8 Appendix

### 8.1 Proofs

#### Proof of Theorem 4.1.

For simplicity, we assume each *subgroup* has the same number of individuals. Once the subgroup is tested to be positive, the number of tests implemented for each subgroup (in the third stage) is a constant. We also assume the population size is large enough. In a group with *k*^*\**^ subgroups, the average number of tests performed for FEAT1 in the first and the second stage is given by:

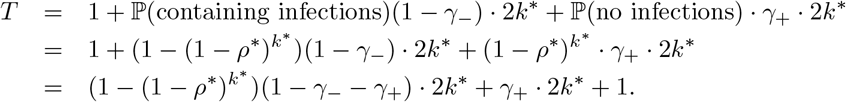

An optimal 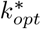 is selected to minimize the average number of tests per subgroup *T/k*^*\**^, i.e.

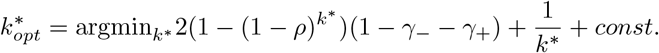

Since 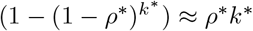 for small *ρ*^*\**^ by Taylor expansion, we aim to find

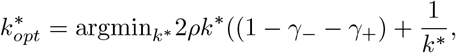

which gives 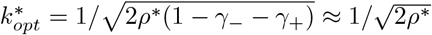 when *γ*_−_, *γ*_+_ are small.□

#### Proof of Theorem 4.2.

Similar to the proof of theorem 4.1, the average number of tests performed for FEAT2 in the first and the second stage of a group is given by:

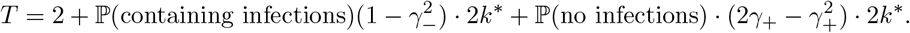

The optimal number of subgroups for each group 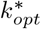 is given by:

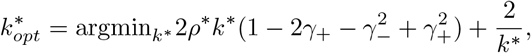

which gives 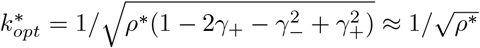 when *γ*_−_, *γ*_+_ are small.□

#### Proof of Theorem 4.3.

For FEAT1, the expected number of tests taken in a group is given by:

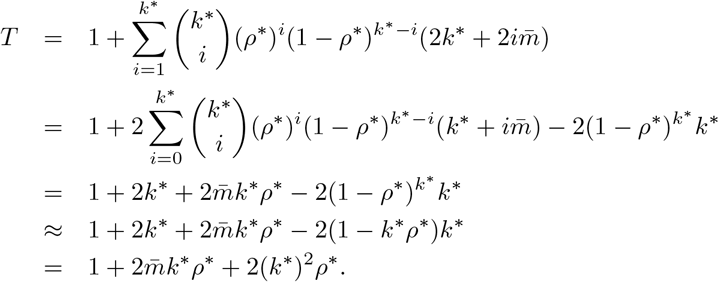

Since a group has 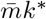 individuals, the expected number of tests per person is given by:

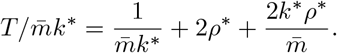

Replace *k*^*\**^ by 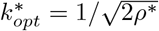,

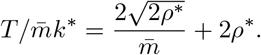

FEAT1 is more efficient than individual testing if and only if 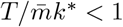, which gives

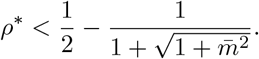

Recall that 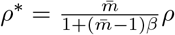, we have

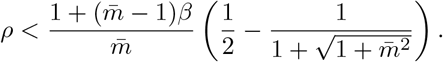

Similarly, FEAT2 is better than individual test if and only if

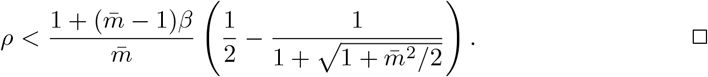

### 8.2 Analysis of group testing accuracy

In this part we provide detailed analysis of the accuracy of group testing strategies to further illustrate the performance in our simulations. When repeated tests are performed, we always apply “once-positive” rule, i.e., a group/subgroup/individual is labeled as positive if either one of the two diagnostic tests is positive.

- **The individual testing**

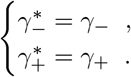

Since the false positive rate and false negative rate are directly determined by the accuracy of a single RT-PCR test.
- **Two-stage strategy with single testing**:

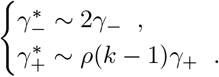

For two-stage strategy like Dorfman’s procedure, each false negative could result from a false negative group testing or, successfully detected in group testing while failed in its individual testing. So 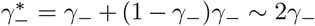. A false positive could come from an infected group or uninfected group. 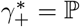 (Healthy, in an infected group)(1−*γ*_−_)*γ*_+_ +𝕡 (Healthy, in an uninfected group) 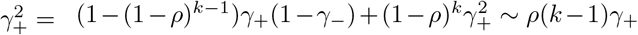 for small *ρ* by Taylor expansion. This explains the simulation results of two-stage methods that 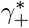 is almost linear in k when *ρ* is small.
- **Three-stage strategy with single testings**:

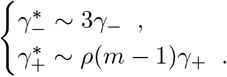

Three-stage strategy with single testings models the case if FEAT does not adopt repeated tests nor infection information in subgroups. For three-stage methods, 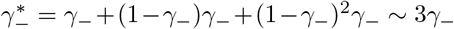. As for false positives, if he comes from a non-infected group, the false positive rate is at order 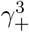. If he comes from a non-infected subgroup and at the same time in an infected group, the false positive rate is at order 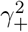. So the decisive order is given by a false positive when he is in an infected subgroup. 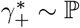 (An healthy person in an infected subgroup) 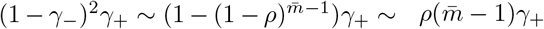. This explained why three-stage procedures have low and stable false positive rates. The false negative rate of group testing strategies without repetition would be doubled or tripled depending on the number of stages, which confirms our simulation results.
- **Two-stage strategy with double testings**:

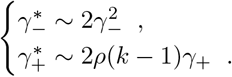

Applying repetition with “once-positive” rule amplifies false positive rate from *γ*_+_ to 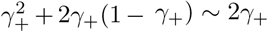, while decrease false negative rate from *γ*_−_ to 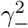 compared with a single dianostic test. For two-stage methods, 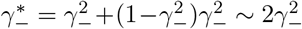, which is controlled far below the false negative rate of widely used individual test at a higher order. 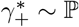 (An healthy person in an infected group) 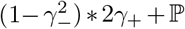 (An healthy person in an uninfected group) 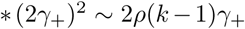 which doubles the false positive rate than before. However, since prevalence *ρ* is small in most cases, false positive rate is well controlled.
- **Three-stage strategy with double testings**:

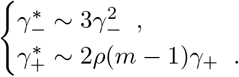

For three-stage strategies with uniform distributed infections, 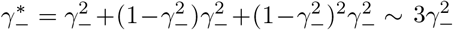. False negative rate of three-stage strategies shrinks to order two. For *γ*_−_ = 0.02 in our cases, three-stage strategies have 1/16 false negative rate compared to the individual test. As for false positives, when false positives appears in an uninfected group or subgroup, false positive rates changed to 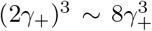 or 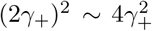 respectively. Even though they are higher than the scenarios when repetitions are not performed, such increasing in false positives is still negligible by their orders. The decisive order of 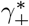 is: 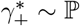 (An healthy person in an infected subgroup) 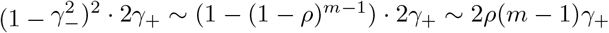
- #### FEAT (proof of theorem 4.4)

For FEAT1:

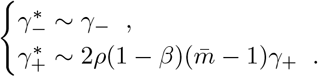

For FEAT2:

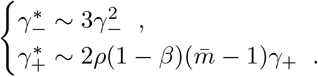

The discussion of the false negative rate of FEAT2 is the same as other three-stage strategies. For FEAT1, 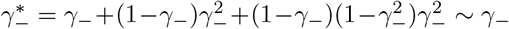. As for the false positive rate, we only need to figure out the probability of an healthy person in a positive subgroup with non-zero infection rate. Once the probability is determined, 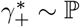 (An healthy person in an infected subgroup)·2*γ*_+_ for both FEAT1 and FEAT2. Here we assume the spread of COVID-19 in the same subgroup are independent conditional on the “source” positive individual.

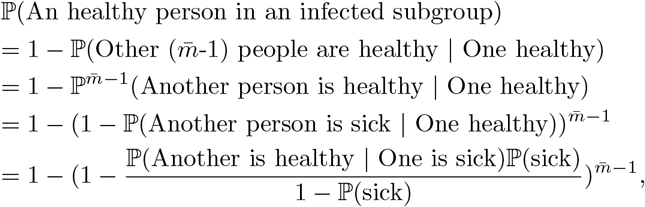

by Bayes theorem.

Here,

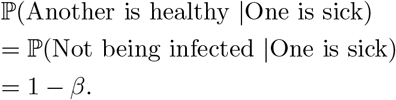

Since *ρ/*(1 − *ρ*) ∼ *ρ*,

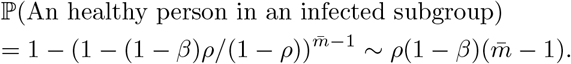

Thus,

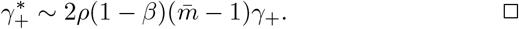

### 8.3 Relationship between *ρ* and *ρ*^*\**^

The relationship between the prevalence *ρ* = 𝕡 (A person is infected) and *ρ*^*\**^ = 𝕡 (A subgroup contains infections) is as follows:

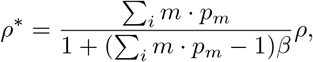

where the size of a subgroup is distributed as 𝕡 (*n*_*i*_ = *m*) = *p*_*m*_, and *β* = 𝕡 (within-subgroup infection rate). The proof of (1) follows from:

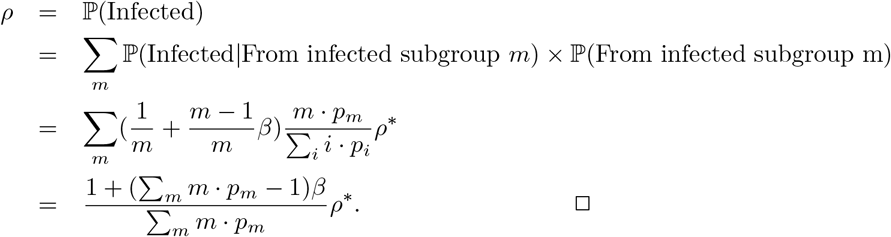

## 9 CODE AVAILABILITY

Our code is available on https://github.com/Fang-Lin93/FAST_group_testing

